# Size Dependent Particle Removal Efficiency and Pressure Drop of a Dust Cleaning Material For Use as Facemask Filters for Protection during COVID-19

**DOI:** 10.1101/2020.07.13.20151985

**Authors:** David Dhanraj, Shruti Choudhary, Pat Raven, Pratim Biswas

## Abstract

COVID-19 pandemic has caused a severe demand for facemasks, and this has resulted in the use of those made from alternate media. As SARS-CoV-2 spreads primarily due to airborne droplets, it is critical to verify the filtration efficiency of these alternate media based facemasks. While several media are being tested and used, commercially available dust cleaners have shown reasonable filtration efficiency. This may also be due to the potential electrostatic charge on the surface which enhances capture of the fine particles. In this manuscript, we report the size dependent filtration efficiency studied systematically in a filter holder-based system as 47 mm punches; and test results on a mannequin that was 3D printed wearing a bandana mask that was placed in a chamber.

## 1. Introduction

COVID-19 pandemic has left numerous communities in a dearth of commercial face masks and thus several masks from alternate filter media are being manufactured (Kampf et al., 2020). SARS-CoV-2 is considered to primarily spread via respiratory droplets (Lu et al., 2020; Wong et al., 2020). SARS-CoV-2 aerosols have increased airborne fitness in comparison with SARS-CoV and MERS-CoV (Fears et al., 2020). Presently, the efficacy of these masks in filtering the virus is unknown. With the governments slowly lifting restrictions, specifically in the United States, several businesses are set to reopen, making it inevitable for individuals to wear face masks. It is important to note that even if these masks are not highly efficient, they will mitigate the spread of the pandemic (Feng et al., 2020).

The size of the SARS-CoV-2 virus is approximately 100 nm in dry state (Zhu et al., 2020), whereas when suspended in respiratory droplets, will be higher and the droplet size can go up to 10 μm or even larger. However, larger droplets settle faster, while smaller droplets have higher airborne lifetime thereby increasing the potential for infection (Biswas & Dhawan, 2020). It is therefore important to evaluate size dependent aerosol filtration efficiency of the alternate filter media, as they are not very efficient. This information will be critical in designing and manufacturing facemasks from readily available, cheap alternate media.

Considering the different materials that are being tested and used, one promising class of materials are dust cleaners such as Swiffer (P&G, Brown et al., 2004). As this is a commonly available material, the objective of this paper was to report the characterization results for Swiffer as an alternate filter media material and evaluate the size-dependent filtration efficiency. Two systems were used: one as 47 mm punches in a filter holder assembly systems, and second when placed on a mannequin as a home-made mask (Bandana type) with swiffer material layered between pillow-case fabrics and in 3-D printed facemask filter holders. The collection efficiency as a function of particle size and the resultant pressure drops are reported.

## 2. Materials and Methods

The filtration efficiency was tested using two different test methods. In the first approach, 47 mm discs were extracted from the filter media and tested in a filter holder assembly, such that the filtration efficiency in a perfectly sealed system can be measured. This would be representative of the efficacy of filtration of the media. While in the second approach, the material was placed in a facemask that was fitted to a mannequin housed in a chamber. These second group of tests are reflective of the filtration efficiency under more realistic conditions accounting for leaks when used by a person. As stated earlier, the dust cleaner material used as filtration media in this study was a Swiffer type material (P&G, Brown et al., 2004). This material has certain electrostatic characteristics to promote the sticking of the dust particles onto the fiber surface.

In both the approaches, the challenge aerosol was generated from a 0.3 M NaCl solution in DI water utilizing a Collision Nebulizer (Single Jet, BGI/Mesa Labs), which was then dried in a Diffusion Drier (Model 3062, TSI Inc.) and then neutralized using a neutralizer, which uses a radioactive source (Kr-85). The neutralized polydisperse challenge aerosols is diluted with Nitrogen gas to achieve the desired number concentration and flow rate of aerosols. In both methods, the Particle Size Distribution (PSD) of the aerosol was measured using a scanning mobility particle sizer (SMPS), upstream and downstream to the filter media assembly. The SMPS consists of an in-line neutralizer, a differential mobility analyzer (TSI DMA Model 3081), in which the particles are classified based on their electrical mobilities, and a condensation particle counter (TSI CPC Model 3750) to count the total number concentration of the classified particles.

### 2.1. Filter holder-based system

The schematic of the filter holder-based system is shown in Figure 1. The diluted challenge aerosols are fed to the filter holder (PN 2220 47mm stainless steel filter holder, Gelman Sciences). A magnehelic differential pressure gauge (Dwyer, Michigan City, IN) was used to measure the pressure drop across the filter holder. The total flow across the filter holder is controlled by a mass flow controller (Omega), downstream the filter holder. The filtration efficiency *(η*_*fe*_) for each size class was calculated as:

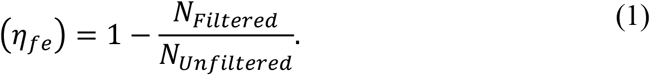

**Figure 1.**
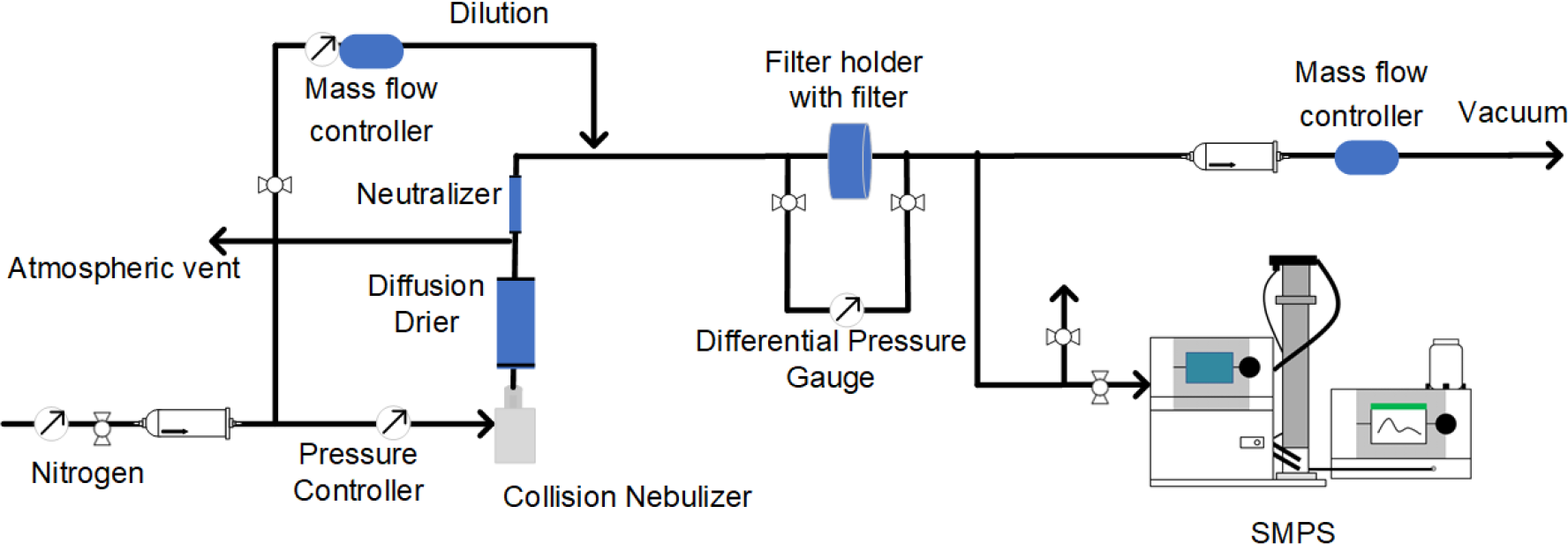
Schematic for filter holder-based system for evaluating size dependent filtration efficiency.

*N*_*filtered*_ and *N*_*UnFiltered*_ were the concentration of each size class measured using the SMPS downstream to the filter holder with and without the filter media.

### 2.2. Mannequin in chamber-based system

The schematic of this method is show in Figure 2. The diluted challenge aerosol was fed into a custom-built acrylic glass chamber (Dimensions: 48 × 24 × 24 inch), sealed with gaskets to ensure uniform size distributions. Two fans were installed at diagonally opposite positions to improve circulation and attain uniform PSD at the sampling points. The facemask was fitted to a mannequin, which was placed at the center of the chamber. The filtration efficiency for each size class was calculated using Eq. 1 wherein *N*_*UnFiltered*_ and *N*_*Filtered*_ were the concentration of each size class measured using the SMPS in front of the masks, and behind the mask though the nostril of the mannequin.

**Figure 2.**
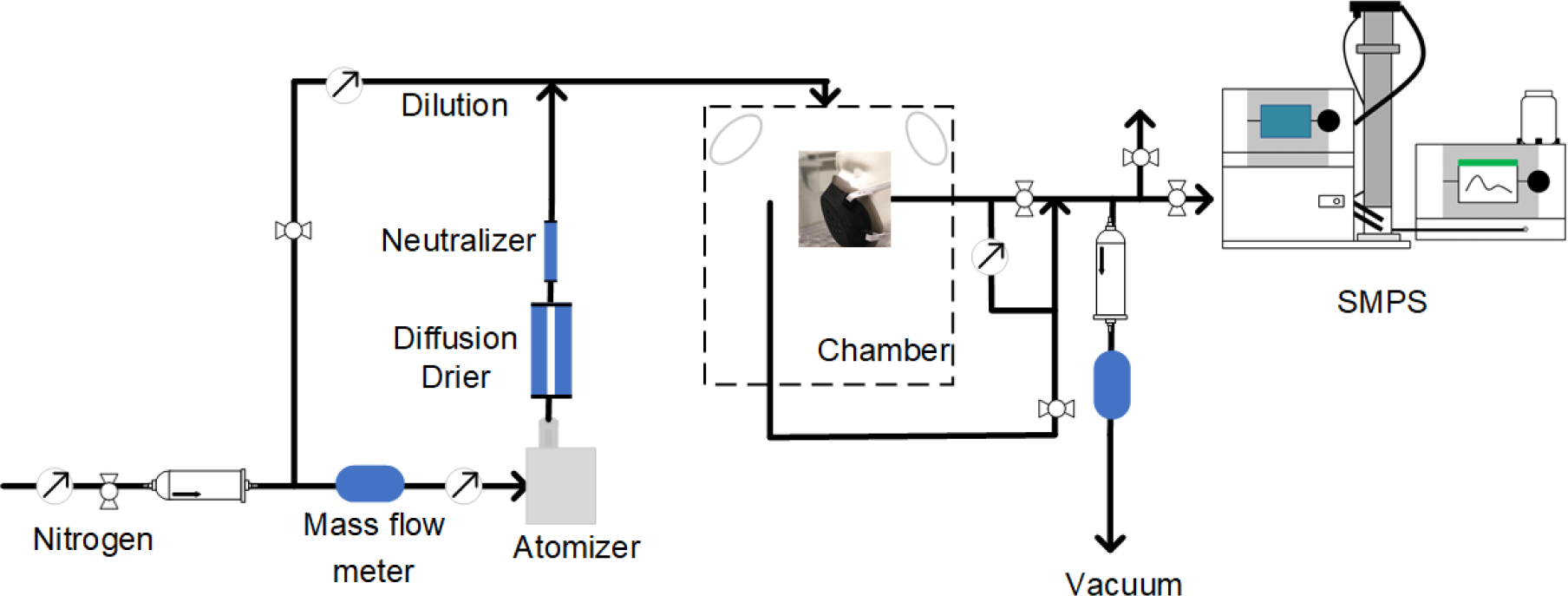
Schematic for mannequin in chamber-based system for evaluating size dependent removal efficiency.

## 3. Results and Discussion

### 3.1. Experimental plan

The list of experiments performed are shown in Table 1. The size dependent filtration efficiency of the Swiffer material was evaluated in the filter holder, 3D printed, and Bandana facemasks. The operating conditions are listed in Table 1.

**Table 1.** Experimental plan

| Section | Filter media |  | Conditions |
| --- | --- | --- | --- |
| Filter holder-based system | Swiffer, Swiffer (2x) and Swiffer (2x) stacked between pillowcase material | | Face velocity: 4.2 cm/s<br>Flow rate: 4.3 Lpm<br>Filter size: 47 mm<br>0.3 M NaCl<br>$N_{Total}: 2 \times 10^6 \text{ \#/cm}^3$<br>11 % RH |
| Mannequin-based system | Mask | Filter media | Face velocity: 4.2 cm/s<br>Flow rate: 4.3 Lpm<br>Exposed filter size: 40 mm<br>0.3 M NaCl<br>$N_{Total}: 1 \times 10^6 \text{ \#/cm}^3$<br>35 % RH |
|  | 3D printed mask | Swiffer(2x) |  |
| | Bandana mask (Sewn using pillowcase fabric) | Swiffer (2x) | Flow rate: 4.3 Lpm, 7 Lpm<br>0.3 M NaCl<br>$N_{Total}: 1 \times 10^6 \text{ \#/cm}^3$<br>28 % RH |

### 3.2. Size dependent removal efficiency estimated in a filter holder-based system

The measured size dependent filtration efficiency of Swiffer punches is shown in Figure 3 (a). The Swiffer material is very efficient in filtering ultra-fine particles (< 80 nm). This is primarily because of the fibrous structures of the Swiffer material trap the ultrafine particles by diffusional capture. The filtration efficiency drops as the particle size increase which is because the fibers are loosely packed and particles that follow the streamlines of the gas get through the material. The highest efficiencies are seen for the case in which two layers of Swiffer is stacked in between two layers of pillowcase fabric. The efficiency ranged from 45 – 62 % for particle sizes between 100 – 500 nm, with the minimum around 300 nm. However, when the layers are doubled, the filtration efficiency increases, and the percent increase is higher for larger particles. For instance, the efficiency of filtration for 200 nm particles increases from 45 % to 65 % (44.4 % increase), whereas for 500 nm, the efficiency increases from 30 % to 58 % (93.3 % increase). The pressure drop for a single layer was 0.01 inch of water and it doubled when the two layers were used, and quadrupled for the third case considered as is shown in Figure 3 (b).

**Figure 3.**
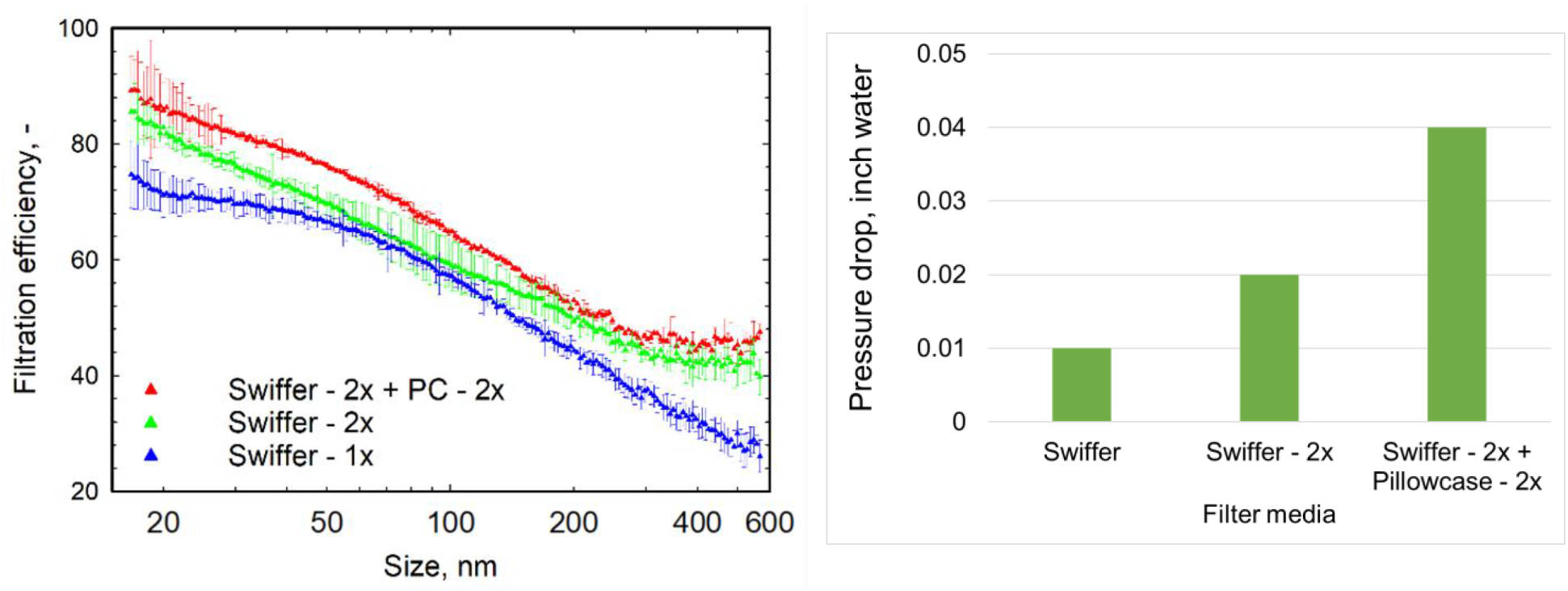
(a) Size dependent filtration efficiency and (b) Pressure drop of Swiffer material tested as single-, double-layer, and double-layer stacked in two layers of pillowcase (PC) 47 mm punches measured in a filter holder-based system.

### 3.3. Size dependent removal efficiency estimated in a mannequin in chamber-based system

#### 3.3.1. 3D printed mask

The 3D printed facemask is shown in Figure 4 (a). The filtration efficiency of a double-layered Swiffer material in the 3D printed face mask and filter holder are shown in Figure 4 (b). Similar observations in the variation of filtration efficiency with particle size can be observed. The filtration efficiency is higher at lower particle sizes and decreases gradually as the size increases. The relative higher filtration efficiency of the smaller particles (< 30 nm) can be attributed to diffusional losses behind the mask. These diffusing particles escape the sampling probe and deposit on the mannequin’s surface. The relative lower filtration efficiency of the larger particles is due to the leakage around the sides on the filter in the 3D printed mask. As larger particles follow the streamlines of the gas, they escape the filter and reach the nostril. It is therefore important to ensure a perfect seal to achieve the highest possible efficiency.

**Figure 4.**
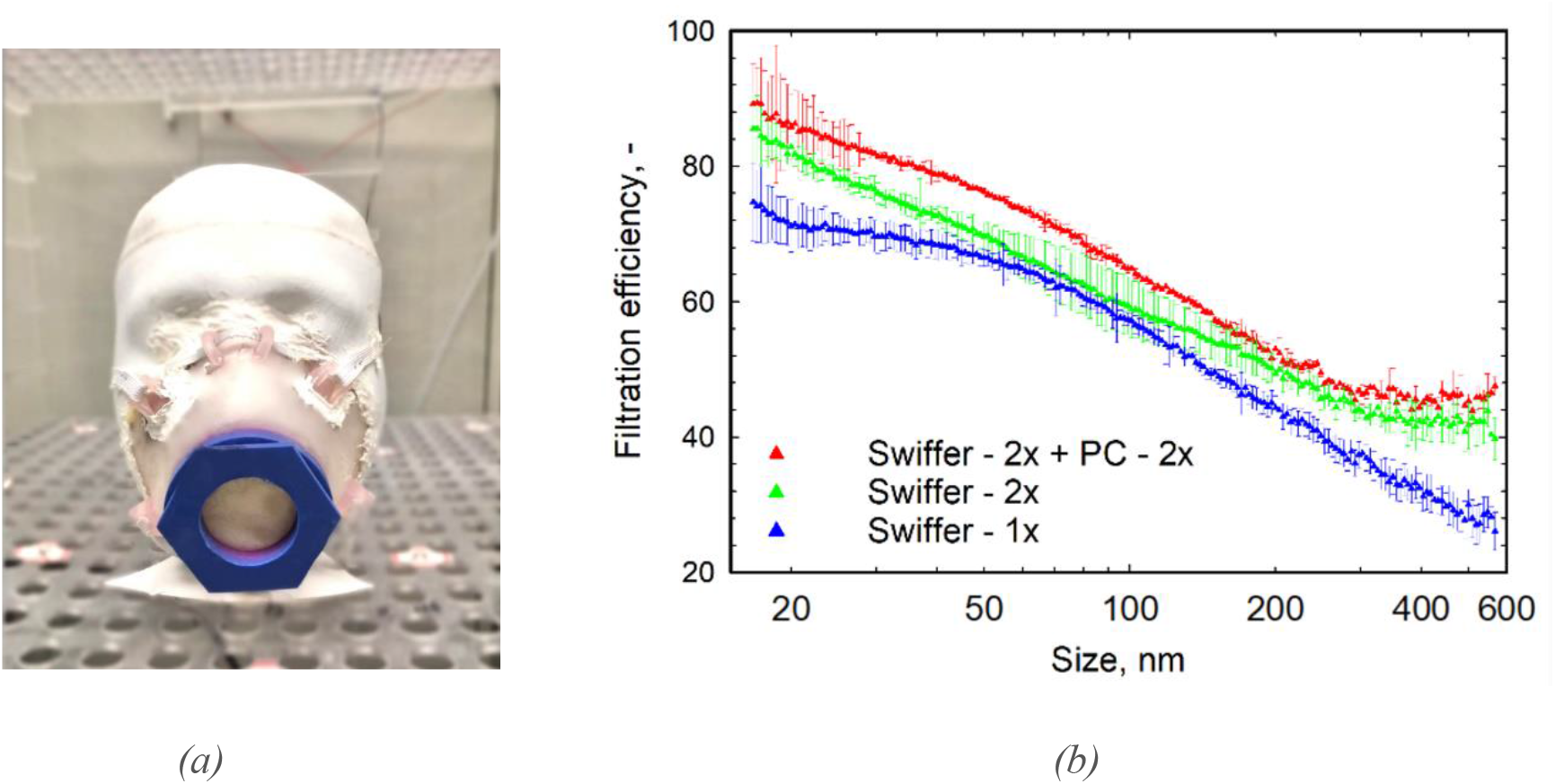
(a) 3D printed face mask, fitted and sealed to mannequin. (b) Size dependent filtration efficiency of Swiffer material tested in a 3D printed facemask compared to the efficiency in a filter holder assembly.

#### 3.2.2. Bandana mask

A fabric-based facemask (Bandana mask) sewn from pillow-case fabric, shown in Figure 5 (a) was tested in the mannequin in chamber-based system with the Swiffer filter material. Two layers of Swiffer material were placed inside the sewn fabric. Its size dependent filtration efficiency as compared to the filtration efficiency measured in a filter holder-based system is shown in Figure 5 (b). The relative higher efficiency of capture of smaller particles in the bandana mask is attributed to diffusional losses during sampling as previously explained in Section 3.2. The filtration efficiency is relatively lower in the Bandana mask as compared to that in a filter holder.

**Figure 5.**
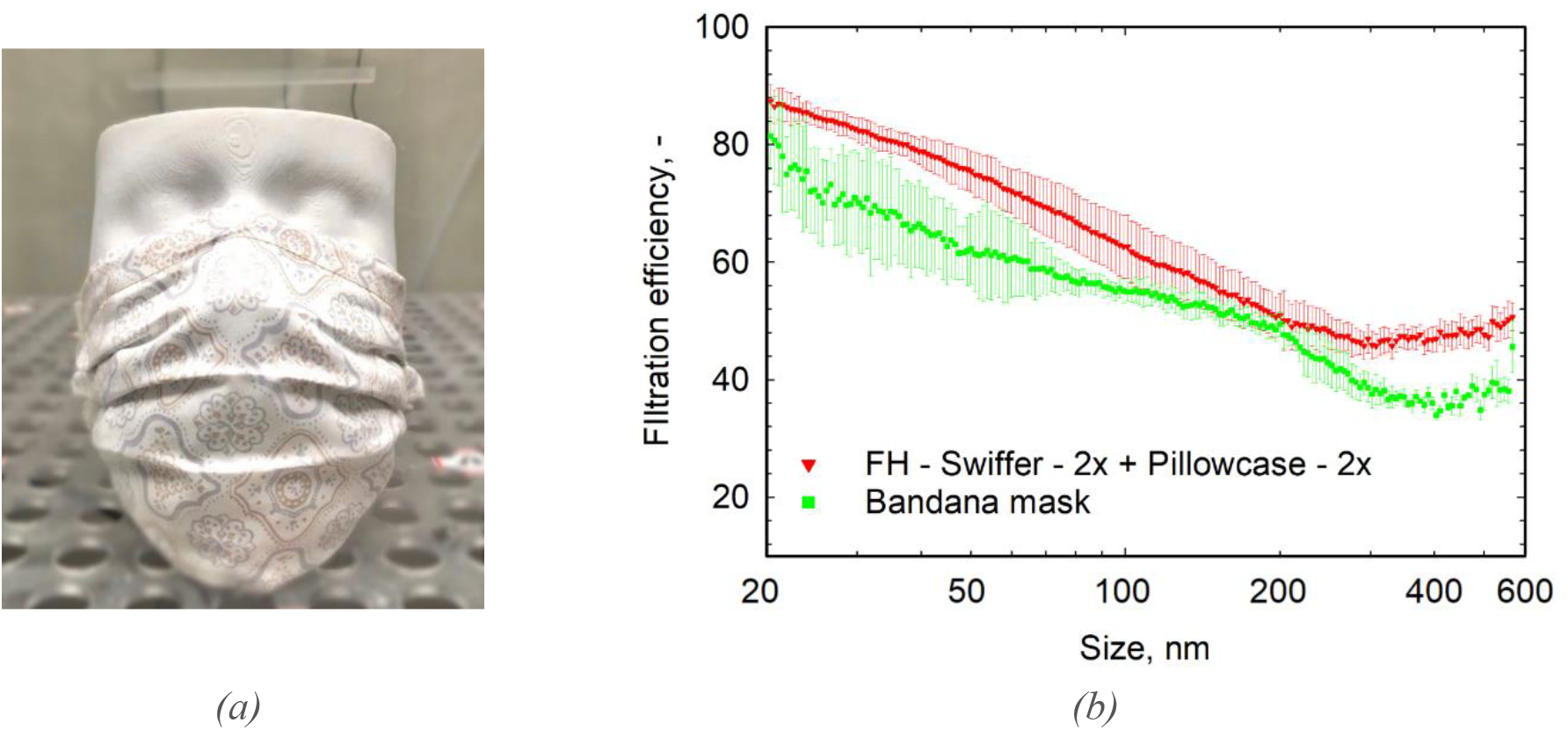
(a) Bandana mask, sewn from pillowcase fabric fitted to mannequin. (b) Size dependent filtration efficiency of Swiffer material tested in Bandana mask compared to its efficiency in the filter holder system. (PC – Pillowcase)

### 3.4. Effect of air inspiration rate and exposure time on size dependent filtration efficiency

The effect of air inspiration rate on filtration efficiency of the Swiffer material in a Bandana mask is studied. The measured size dependent filtration efficiency is shown in Figure 6 (a). The filtration efficiency drops for higher inspiration rate and the extent of decrease is higher for larger particles as compared to smaller particles. Furthermore, Figure 6 (b) shows the effect of filter exposure time on a double layer 47 mm punch in the filter holder system. Each SMPS scan takes a minute and hence the filtration efficiency at the first, 12^th^ and 18^th^ minutes are shown. While, the variation in efficiency is not significant, it can be observed that the filtration efficiency increases with time for smaller particles and decreases with time for larger particles.

**Figure 6.**
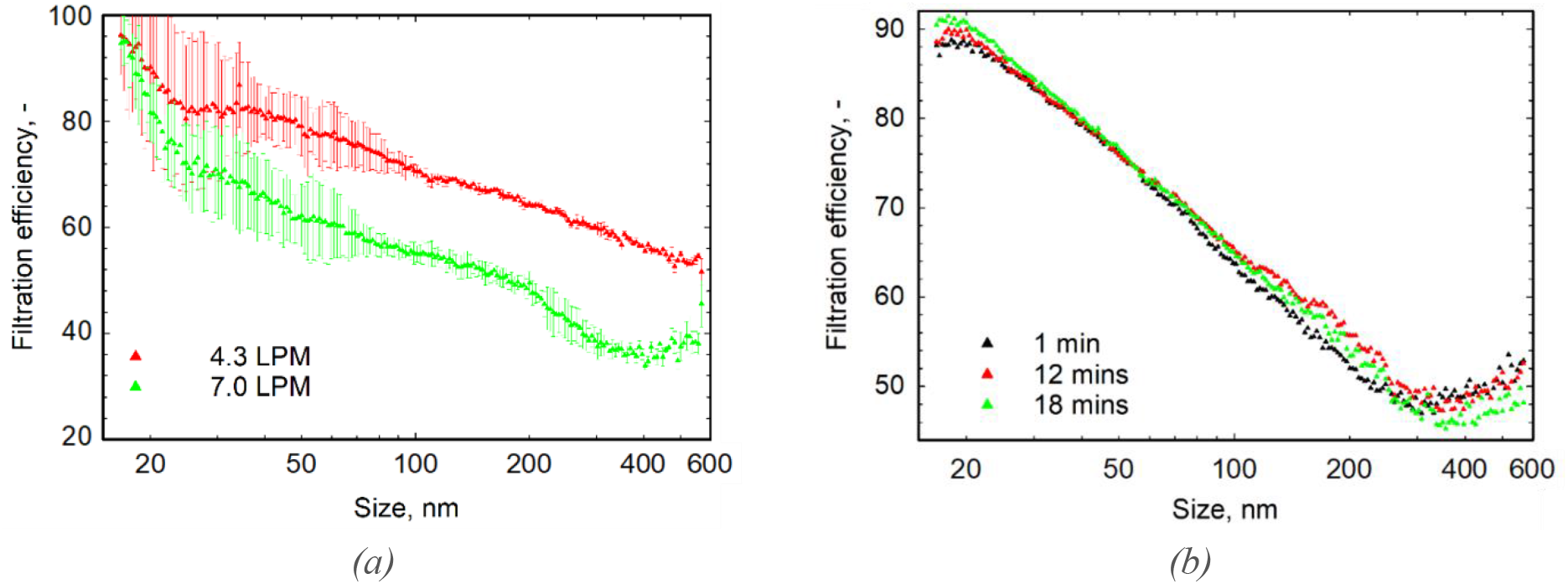
Effects of (a) inspiration flowrate and (b) exposure time on size dependent filtration efficiency of Swiffer material.

## 4. Conclusions

The size dependent filtration efficiency of dust cleaning swiffer material in a filter holder- and mannequin in chamber-based systems is reported. It was found that the highest filtration efficiencies for sizes between 100 – 500 nm, ranged between 45 – 62 % with a minimum at 300 nm for the case in which two layers of Swiffer were stacked within two layers of pillowcase fabric tested in the filter holder-based system. The filtration efficiency under these conditions when tested on 3D printed mannequins and masks were close to the filter holder-based system and was relatively lower in the bandana type mask.

## Data Availability

All data is available from PhD student laboratory notebooks

## 5. Acknowledgement

The authors would like to thank David Ballard and Uday Jammalamadaka from Mallinckrodt Institute of Radiology, Washington University School of Medicine, for designing and making the 3D printed mask. Detailed results from a series of studies collaboratively are being prepared for publication.

